# Evaluation of aortic valve calcification in patients with suspected cardiac amyloidosis

**DOI:** 10.1101/2024.01.09.24301081

**Authors:** Daniel Ng, Karine Moussa, Francis W. Allinson, Adam A. Behroozian, Timothy M. Jordan, Leah Puglisi, Curtiss T. Stinis, Paul S. Teirstein, Rajeev C. Mohan

## Abstract

**Background:** The association between transthyretin cardiac amyloidosis (TTR-CA) and aortic stenosis (AS) has been described, although the mechanism by which they interact remains unclear. It has previously been proposed that this may be secondary to excess valvular calcification.

**Objectives:** We propose that patients with suspected cardiac amyloidosis will have increased aortic valve calcification, as evidenced by elevated aortic valve calcium score on CT imaging.

**Methods:** We retrospectively identified patients with severe AS referred for transcatheter aortic valve implantation (TAVI) from January 2017 to November 2022. This population was divided into two cohorts; the Likely CA cohort had suspected TTR-CA by echocardiogram while the Unlikely CA cohort did not. Baseline characteristics, echocardiographic data, CT aortic valve calcium scores, and post-procedural complications were compared.

**Results:** Of the 496 patients analyzed, 145 (29.2%) patients met echocardiographic criteria (interventricular septal thickness (IVS) ≥1.2 cm and average mitral annular systolic s’ ≤ 6 cm/s) for the Likely CA cohort. The Likely CA cohort was more likely to be older, be male, have atrial fibrillation, and have worse renal function. On echocardiogram, the Likely CA cohort had increased hypertrophy, worsened diastolic function, and decreased systolic function. Aortic valve calcium score by CT was not significantly different between the Likely CA and Unlikely CA cohort (2834.95 AU compared to 2852.27 AU, p=0.914). There was no statistically significant difference in post-TAVI complications.

**Conclusions:** Patients with an echocardiographic profile consistent with TTR-CA had no difference in aortic valve calcification in comparison to a population unlikely to have TTR-CA.

## Introduction

Cardiac amyloidosis (CA) is characterized by the deposition of misfolded proteins, or amyloid fibrils, within the myocardium. Transthyretin (TTR) is one of the most common proteins implicated in CA^1^. TTR-CA is the root of multiple chronic cardiac conditions, including diastolic dysfunction, atrial fibrillation, and aortic stenosis (AS)^2^. There are several echocardiographic features of TTR-CA that have been previously described, including myocardial hypertrophy, small left ventricular chamber size, biatrial enlargement, valvular thickening, impaired diastology, reduced atrial function, and reduced systolic function as evidenced by global longitudinal strain^3^. The echocardiographic assessment of AS is most notable for elevated valvular velocities and gradients^4^. However, there are other echocardiographic findings in patients with severe AS that overlap with TTR-CA, such as left ventricular hypertrophy^5^, impaired global longitudinal strain^6^, and diastolic dysfunction^7^. However, the degree of hypertrophy is significantly greater in patients with TTR-CA related AS compared to lone AS^8^. Systolic dysfunction is also diminished to a greater degree in TTR-CA patients with severe AS, as evidenced by average mitral annular s’ and global longitudinal strain^8^.

While these echocardiographic findings are helpful in identifying patients with TTR-CA and severe AS, they do not explain the relationship between the two conditions. One hypothesis suggests that amyloid directly deposits into the valve leaflets, causing abnormal thickening and eventual AS^9^. Conversely, there has been histologic evidence in explanted stenotic aortic valves of colocalization between amyloid and calcific deposits, which suggests that amyloidosis may promote abnormal calcification^10^. This has been contested by a later study that showed TTR-CA patients had significantly decreased aortic valve calcification on CT^11^. As such, the mechanism by which TTR-CA associated AS develops remains unknown. We propose patients with severe AS being evaluated for transcatheter aortic valve implantation (TAVI) and suspected TTR-CA by echocardiogram will have increased aortic valve calcification. The combination of average mitral annular s’ velocity ≤6 cm/s and interventricular septum (IVS) ≥1.2 cm to identify patients with likely TTR-CA has been previously described in an analysis of the TOPCAT trial^12^.

## Methods

This was a single-center, retrospective analysis of patients with severe AS referred for TAVI at Scripps Memorial Hospital in San Diego, California between January 2017 and November 2022. The design of this study was approved by the Scripps Health Institutional Review Board and Ethics Committee and performed in accordance with institutional guidelines. A total of 1053 patients met the inclusion criteria of severe aortic stenosis. Patients with prior aortic valve replacement, non-tricuspid aortic valve morphology, or missing data were excluded. After exclusions, 496 patients remained available for analysis. Baseline characteristics were obtained from the STS/ACC TVT registry.

Echocardiograms used for analysis were performed within one month prior to TAVI or within 2 days post-TAVI within the Scripps Health system. Echocardiograms were performed by sonographers using Philips EPIQ, Vivid E95, or Philips IE33 ultrasound machines. Global longitudinal strain (GLS) was obtained only on echocardiograms with sufficient endomyocardial border visualization.

Patients were divided into two cohorts. The enriched (Likely CA) cohort consisted of patients with both IVS ≥1.2 cm and average mitral annular systolic s’ ≤ 6 cm/s. The unenriched (Unlikely CA) cohort met only one or none of the echocardiographic criteria.

The primary outcome was aortic valve calcium score by CT imaging. Secondary outcomes included post-procedural complications, defined as post-procedural atrial fibrillation, stroke or transient ischemic attack, need for pacemaker, bleeding, vascular complications, or death.

To discern demographic and clinical characteristic differences between groups, all categorical variables were summarized as frequencies and percentages and compared using the Chi-Square test or Fisher’s exact test. All continuous variables were summarized as means and standard deviations and compared by t-tests if normally distributed. If not normally distributed, they were summarized as medians and interquartile ranges and tested by Mann-Whitney U tests. All analyses were conducted using R v. 4.1.3 and/or SPSS v28, and p-values <0.05 were considered statistically significant.

## Results

Of the 496 patients analyzed, 145 (29.2%) patients met criteria for the Likely CA cohort and the remaining 351(70.8%) patients were in the Unlikely CA cohort. Baseline demographics are shown in **Table 1**. The Likely CA cohort was noted to be significantly older (82.72 years compared to 79.95 years, p<0.001), male-predominant (69.0% compared to 57.8%, p=0.021), higher prevalence of atrial fibrillation (49.7% compared to 31.9%, p<0.001), and have higher creatinine (1.1 compared to 0.9, p=0.009). There was no statistically significant difference in the primary end point of aortic valve calcium score between the Likely CA and Unlikely CA cohort (2834.95 AU compared to 2852.27 AU, p=0.914).

**Table 1:** Baseline demographics.

|  | **Unlikely CA cohort**<br>**(N=351)** | **Likely CA cohort**<br>**(N=145)** | **p-value** |
| --- | --- | --- | --- |
| **Age (years)** | 79.95 ± 9.17 | 82.72 ± 7.63 | <0.001 |
| **Male** | 203 (57.8) | 100 (69.0) | 0.021 |
| **White** | 332 (94.6) | 132 (91.0) | 0.143 |
| **Coronary artery disease** | 175 (49.9) | 84 (57.9) | 0.102 |
| **Hypertension** | 308 (87.8) | 130 (89.7) | 0.548 |
| **Heart failure** | 271 (77.2) | 117 (80.7) | 0.393 |
| **Atrial fibrillation** | 112 (31.9) | 72 (49.7) | <0.001 |
| **Diabetes** | 110 (31.3) | 43 (29.7) | 0.712 |
| **NYHA classification** |  |  |  |
| **None** | 15 (4.3) | 4 (2.8) | 0.051 |
| **Class I** | 19 (5.4) | 2 (1.4) |  |
| **Class II** | 130 (37.0) | 51 (35.2) |  |
| **Class III** | 176 (50.1) | 77 (53.1) |  |
| **Class IV** | 11 (3.1) | 11 (7.6) |  |
| **KCCQ12 score** | 62.69 ± 27.08 | 62.13 ± 27.34 | 0.848 |
| **Creatinine (mg/dL)** | 0.905 ± 0.473 | 1.1 ± 0.65 | 0.009 |
| **AV calcium score (AU)** | 2852.27 ± 1692.35 | 2834.95 ± 1602.25 | 0.914 |

Values are expressed as mean ± SD or n (%). Creatinine is expressed as median ± IQR. Likely CA cohort was defined as mitral annular s' velocity ≤6 cm/s and interventricular septal thickness ≥ 1.2 cm.

Echocardiographic data was compared between the two cohorts (**Table 2**). The Likely CA cohort had significantly decreased left ventricular ejection fraction (LVEF), increased IVS thickness, increased left ventricular posterior wall thickness, decreased left ventricular internal diameter, increased left ventricular mass, increased E/e’ ratio, decreased average mitral annular s’ velocity, and decreased stroke volume. There was no statistically significant difference in left atrial volume (p=0.572). Longitudinal strain was compared between the two cohorts (**Table 3**). The Likely CA cohort had significantly reduced average GLS, but there was no difference in relative apical longitudinal strain index (RapLSI).

**Table 2:** Echocardiographic parameters.

|  | **Unlikely CA cohort**<br>**(N=351)** | **Likely CA cohort**<br>**(N=145)** | **p-value** |
| --- | --- | --- | --- |
| **LVEF (%)** | 60.8% ± 12.4% | 55.2% ± 12.9% | <0.001 |
| **IVS (cm)** | 1.16 ± 0.23 | 1.43 ± 0.22 | <0.001 |
| **LVPWd (cm)** | 1.1 ± 0.20 | 1.24 ± 0.19 | <0.001 |
| **LVIDd (cm)** | 4.60 ± 0.63 | 4.56 ± 0.71 | <0.001 |
| **LAVI (mL/m<sup>2</sup>)** | 40.57 ± 15.20 | 46.29 ± 15.33 | 0.572 |
| **LV mass (g)** | 192.83 ± 62.16 | 241.64 ± 72.15 | <0.001 |
| **E/e' ratio** | 14.9 ± 8.5 | 18.8 ± 12.5 | <0.001 |
| **Average mitral annular s' velocity (cm/s)** | 6.58 ± 1.79 | 5.17 ± 1.22 | <0.001 |
| **Stroke volume (mL)** | 86.43 ± 28.72 | 74.56 ± 25.20 | <0.001 |

Values are expressed as mean ± SD. E/e' ratio and average mitral annular s' are expressed as median ± IQR. IVS = interventricular septal thickness. LAVI = left atrial volume index. LVEF = left ventricular ejection fraction. LVIDd = left ventricular internal diameter in end diastole. LV mass = left ventricular mass. LVPWd = left ventricular posterior wall thickness in end diastole.

**Table 3:**
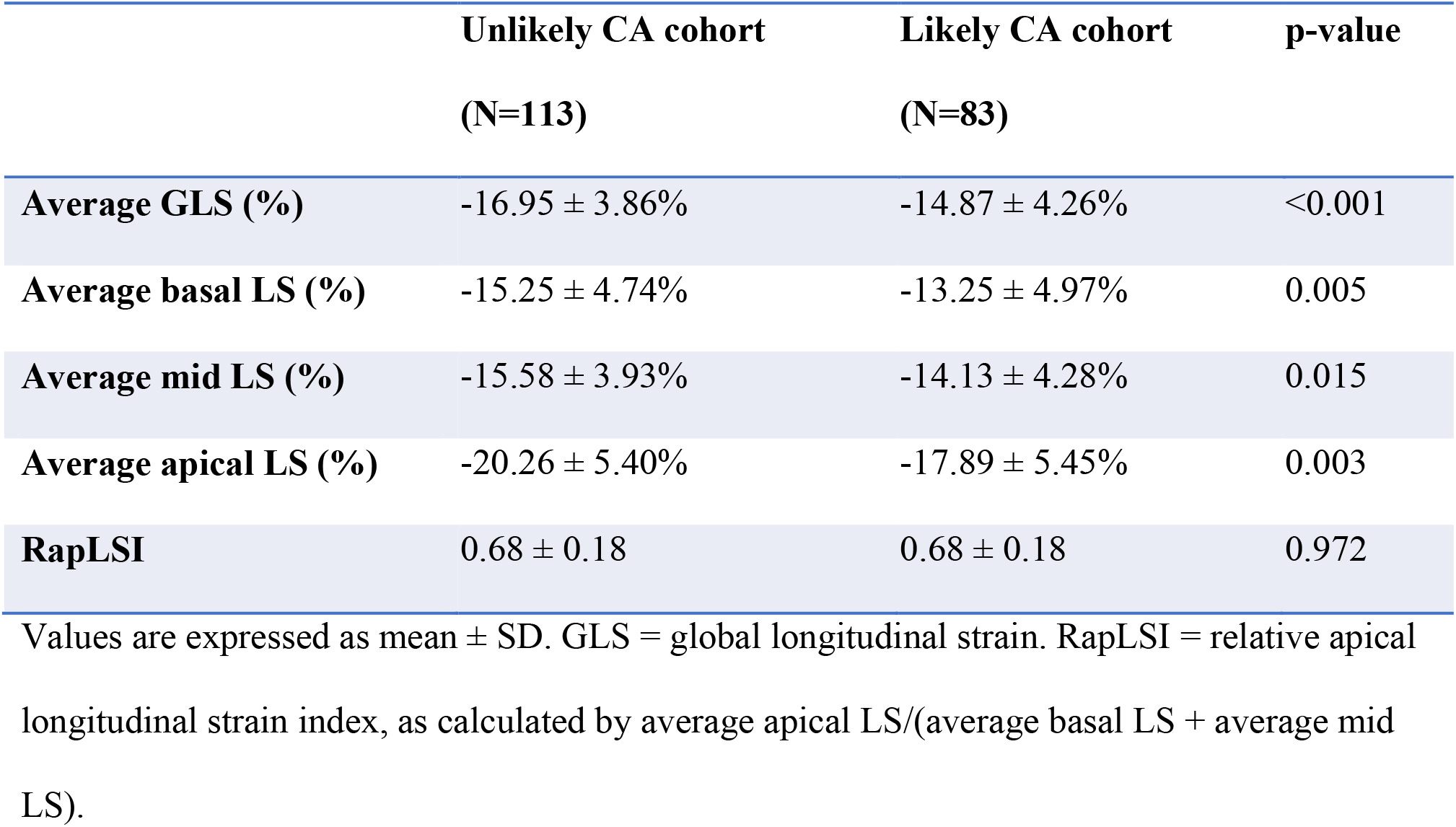
Global longitudinal strain.

There was a non-significant trend towards an increased rate of post-procedural atrial fibrillation in the Likely CA cohort compared to the Unlikely CA cohort (p=0.051). Otherwise, there was no significant difference in other post-procedural endpoints including stroke or transient ischemic attack, need for pacemaker, bleeding, vascular complications, or death **(Table 4)**.

**Table 4:** Post-TAVI complications.

|  | **Unlikely CA cohort**<br>**(N=351)** | **Likely CA cohort**<br>**(N=145)** | **p-value** |
| --- | --- | --- | --- |
| **Atrial fibrillation** | 3 (0.9) | 5 (3.5) | 0.051 |
| **CVA/TIA** | 4 (1.1) | 4 (2.8) | 0.240 |
| **Pacemaker** | 17 (4.8) | 6 (4.1) | 0.819 |
| **Bleed** | 7 (2.0) | 4 (2.8) | 0.738 |
| **Vascular complication** | 12 (3.4) | 4 (2.8) | >0.999 |
| **Death** | 2 (0.6) | 2 (1.4) | 0.584 |

Values are expressed as n (%). CVA = cerebrovascular event. TAVI = transcatheter aortic valve implantation. TIA = transient ischemic attack.

## Discussion

In our population of patients with severe AS, 29.2% met echocardiographic criteria that was suspicious for TTR-CA, which is higher than a prior reported estimate of 15-16%^8,13^. The Likely CA cohort was older, which is concordant with the current literature on the epidemiology of TTR-CA^14^. There was a significantly higher prevalence of atrial fibrillation in the Likely CA cohort, in line with the known association between TTR-CA and atrial fibrillation^15^.

The Likely CA cohort displays the same echocardiographic phenotype as that of TTR-CA, including left ventricular hypertrophy, impaired diastolic function, and reduced left ventricular systolic function. This is concordant with recent data that demonstrates that TTR-CA can still be identified by echocardiogram even in the presence of AS^16^. The Likely CA cohort had significantly reduced GLS but no difference in RapLSI. It has been previously described that severe AS on its own may mimic the relative apical sparing pattern^17^, which may explain the unchanged strain pattern between our two cohorts. Secondly, the apical sparing pattern may depend on disease progression, as a prior study noted that only half of patients with proven TTR-CA had the pattern at the time of diagnosis^18^.

The primary endpoint of aortic valve calcium score by CT was similar between the Likely CA and Unlikely CA cohorts. Our findings suggest that calcific aortic valve disease is not the sole driver for the development of AS in TTR-CA patients. Further studies are needed to assess the mechanism by which TTR-CA promotes AS.

TTR-CA patients with severe AS benefit from TAVI, with significant improvement in survival compared to medical management^1^. Recently, there has been focus on targeted drug therapy for TTR-CA. The ATTR-ACT trial found that tafamidis reduced all-cause mortality and improved quality of life by stabilizing transthyretin and reducing tissue deposition^20^. The impact of tafamidis on AS in TTR-CA patients remains unknown.

### Limitations

Our study had two primary limitations. While patients in the Likely CA cohort had echocardiographic findings consistent with TTR-CA, no confirmatory testing with bone scintigraphy or endomyocardial biopsy was performed. Additionally, due to the small sample size, the study was underpowered to assess for post-procedural complications. Lastly, there was a significant proportion of patients who did not have adequate endomyocardial border differentiation for strain analysis.

## Conclusions

Patients with severe AS and likely TTR-CA had no difference in aortic valve calcification compared to patients with lone severe AS, suggesting that TTR-CA promotes AS via a different mechanism.

## Data Availability

The data was obtained from the STS/ACC TVT of Scripps patients as approved by the Scripps IRB.

